# Comparative Transcriptome Analysis of Acne vulgaris, Rosacea, and Hidradenitis Suppurativa Supports High Dose Dietary Zinc as a Therapeutic Agent

**DOI:** 10.1101/2023.11.28.23299152

**Authors:** Li Li, Irshad Hajam, Jean S. McGee, Zhengkuan Tang, Ye Zhang, Nikil Badey, Esther Mintzer, Zhenrui Zhang, George Y. Liu, George M. Church, Yu Wang

## Abstract

Acne vulgaris, rosacea, and hidradenitis suppurativa are enduring inflammatory skin conditions that frequently manifest with akin clinical attributes, posing a considerable challenge for their distinctive diagnosis. While these conditions do exhibit certain resemblances, they also demonstrate distinct underlying pathophysiological mechanisms and treatment modalities. Delving into both the molecular parallels and disparities among these three disorders can yield invaluable insights for refined diagnostics, effective management, and targeted therapeutic interventions. In this report, we present a comparative analysis of transcriptomic data across these three diseases, elucidating differentially expressed genes and enriched pathways specific to each ailment, as well as those shared among them. We also identified high dose dietary zinc as a potential therapeutic agent and validated its efficacy in an acne mouse model.

## INTRODUCTION

Acne vulgaris, Rosacea (formally called Acne rosacea), and Hidradenitis Suppurativa (HS, formally called Acne inversa) are chronic inflammatory skin disorders that can cause substantial physical and psychological distress for patients. While these three disorders are distinct dermatological conditions, they do share some clinical manifestations(Katsambas and Dessinioti 2017). Acne vulgaris is characterized by follicular hyperkeratinization, clogged hair follicles, excess sebum production, bacterial colonization, inflammation, and hormonal influences. It primarily affects sebaceous gland-rich areas including the face, chest, and back. Rosacea, on the other hand, involves facial redness, visible blood vessels, and sometimes papules and pustules. Hidradenitis suppurativa mainly affects areas with apocrine sweat glands, resulting in recurrent abscesses, nodules, and sinus tracts (Katsambas and Dessinioti 2017). The diagnosis of these diseases heavily relies on the visual assessment and symptom evaluation performed by skilled dermatologists. Moreover, it’s not uncommon for individuals to experience a combination of these conditions or for one to pave the way for another, further complicating their differential diagnosis. The potential of misdiagnosis could significantly hamper treatment outcomes (Wang et al. 2015). Furthermore, while various treatment options exist, their efficacy displays variability, and with the exception of isotretinoin, which can confer remission in select cases of acne vulgaris, none furnish an unequivocal remedy. Thus, a more profound comprehension of both shared characteristics and distinctions—especially on a molecular level—would not only lead to more accurate diagnoses but also propel the development of improved treatments and eventual cures.

The application of omics technologies has substantially deepened our grasp of the molecular changes that drives skin disorders. Through a comprehensive multi-omic analysis of acne patients’ lesion and non-lesion skin samples, researchers identified a differential gene expression profile enriched in keratinization, immune response, and lipid metabolism and biosynthesis (Hall et al. 2019). Similarly, a transcriptome analysis of rosacea patients’ lesion samples indicated a Th1/Th17 polarized inflammation profile and significant macrophage infiltration across all rosacea subtypes (Buhl et al. 2015). Likewise, a study on HS showcased varying gene expression in immune response, antimicrobial response, and cell differentiation within follicular and epidermal keratinocytes (Buhl et al. 2015).

Considering the shared and distinct clinical manifestation of acne vulgaris, rosacea and HS, we hypothesized that these diseases may exhibit both disease-shared and disease-specific molecular features in their pathogenesis, which could be investigated by comparing the transcriptome profiles of three diseases. We focused on one rosacea subtype, Papulopastular Rosacea (PPR), due to its clinical likeness to acne vulgaris and HS, comparing to other subtypes of rosacea. We collected three transcriptomic datasets, each for the three diseases from NCBI’s Gene Expression Omnibus (Acne vulgaris: GSE6475; PPR: GSE65914; HS: GSE148027; **Supplementary** Fig. 1a) (Buhl et al. 2015; Hall et al. 2019; Zouboulis et al. 2020). Eight additional datasets (Acne vulgaris: GSE53795, GSE108110; PPR: Dataset 1 and Dataset 2; HS: GSE122592; GSE213761; GSE154773; GSE72702) were also utilized for validation (Kelhälä et al. 2014; Carlavan et al. 2018; Shih et al. 2020; Medgyesi et al. 2020; Witte-Händel et al. 2019; Lowe et al. 2020; Gudjonsson et al. 2020; Blok et al. 2016). **Figure 1a** illustrates the project’s overarching design: conducting bioinformatic analyses on the datasets to identify differentially expressed genes (DEGs), followed by comparing these DEGs and enriched pathways across the three diseases to pinpoint those shared and unique. Subsequently, gene-drug interaction analyses were conducted to identify potential drug candidates targeting highly dysregulated genes, with subsequent validation on animal models to mitigate disease severity.

**Figure 1.**
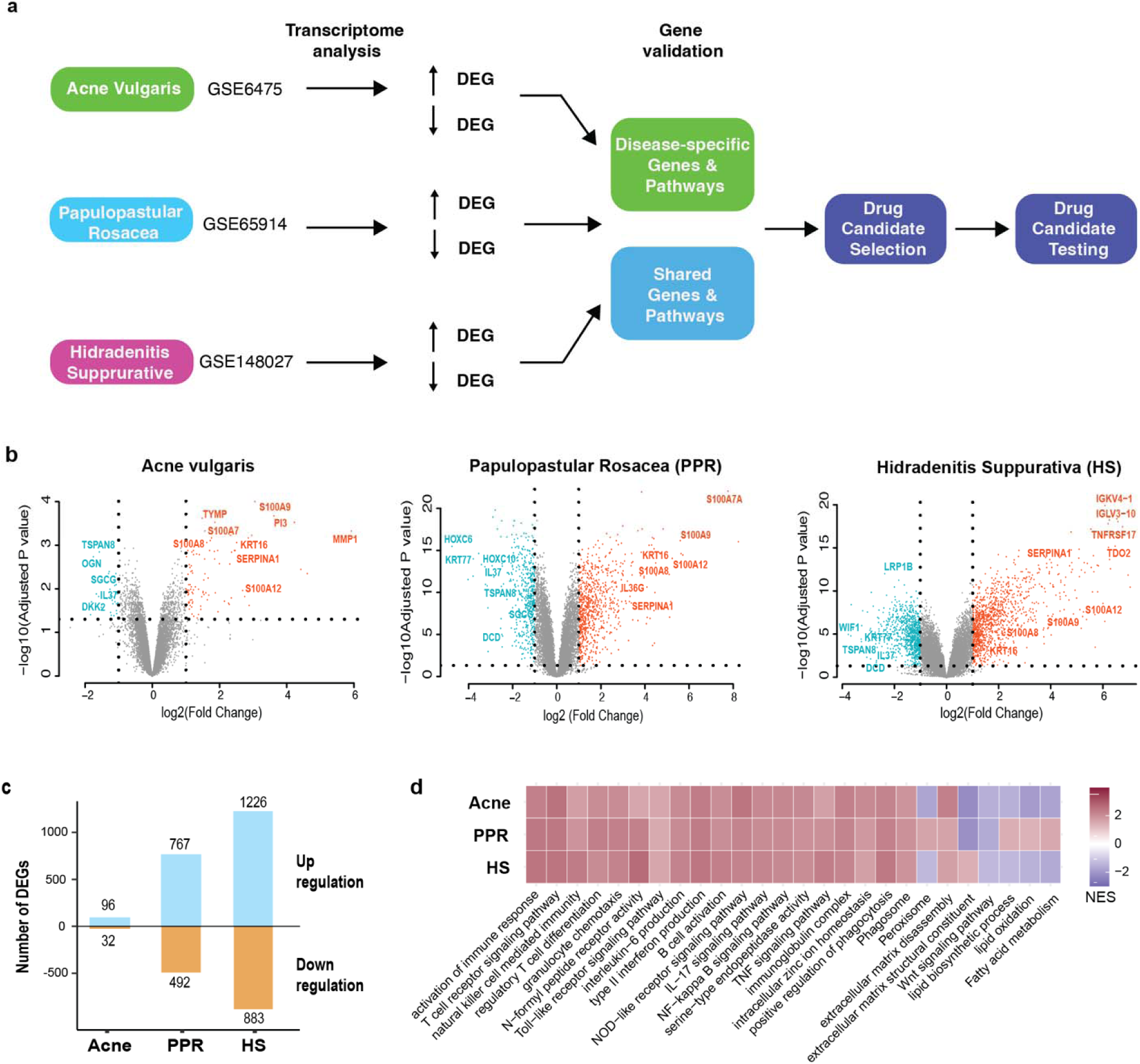
Transcriptome analysis of Acne vulgaris, Papulopastular rosacea (PPR) and Hidradenitis supprurative (HS). **a)** Project overview. Transcriptome data from three main datasets, one for each disease, were analyzed to determine differentially expressed genes (DEGs) and enriched pathways. Disease-specific DEGs an shared DEGs were identified and validated using additional datasets. Drug candidates were screened against selected DEGs and validated on an acne animal model. **b)** Volcano Plots Illustrating DEGs. Genes with a substantial fol change in gene expression and low adjusted P values are prominently highlighted. **c)** Summary of the number of DEGs for each disease. **d)** Selected enriched pathways from gene set enrichment analysis of gene expression profiles for each disease. NES: normalized enrichment score.

## Results

### Identification of differentially expressed genes and enriched pathways through transcriptome analysis

We first conducted differential expression analysis on the eleven datasets (**Fig. 1b and c; Supplementary Fig. 1b and c**), and filtered DEGs using a threshold of adjusted p value [<[0.05 and |log2 Fold Change|[>[1. The examination of the Acne vulgaris dataset (GSE6475) revealed 96 upregulated genes and 32 downregulated genes in lesional samples compared to healthy controls, along with 103 upregulated genes and 9 downregulated genes in lesional samples compared to non-lesional controls. From PPR GSE65914, we identified 767 upregulated genes and 492 downregulated genes in lesional samples when compared with healthy controls. From PPR Dataset 2, 345 upregulated genes and 84 downregulated genes were identified in lesional samples compared to non-lesional controls. In HS GSE148027, our analysis revealed 1226 upregulated genes and 883 downregulated genes in lesional samples compared to healthy controls, as well as 1303 upregulated genes and 823 downregulated genes in lesional samples compared to non-lesional controls. In Acne vulgaris, the most prominently upregulated genes (Lesional versus Healthy) encompassed matrix metalloproteases (MMP1, MMP3, MMP12), members of the serpin family (SERPINA1, SERPINB3, SERPINB4), and genes from the epidermal differentiation complex (SPPR gene family, S100 gene family). Notably, PPR and HS also exhibited substantial upregulation in these genes. Among the most consistently downregulated genes across all three datasets were the motility-promoting protein TSPAN8 and the anti-inflammatory IL-37 in lesional samples compared to healthy controls. This pattern was mirrored in both the PPR and HS datasets, which also demonstrated marked downregulation of the Wnt signaling inhibitor WIF1, the antimicrobial DCD, and the KRT77 genes.

We then conducted gene set enrichment analysis (GSEA) on the three datasets to identify enriched Gene Ontology and KEGG pathways (**Fig. 1d** for lesional versus healthy controls, **Supplementary Fig. 1d** for lesional versus non-lesional controls). All three diseases showed significant upregulation of immune response pathways, such as innate and adaptive immune cell recruitment and activation, associated with regulatory/inhibitory immune responses such as regulatory T cell differentiation. Furthermore, all three diseases demonstrated heightened serine-type endopeptidase activities, leading to increased extracellular matrix disassembly. All three diseases exhibited downregulation in genes involved in the Wnt signaling, especially for genes that possess Wnt inhibitor functions (including WIF1 and DKK2), suggesting a dysregulation of the pathway in the diseases. Both Acne and HS showed downregulation of lipid-related pathways, such as lipid synthesis and modification. PPR showed upregulation in those pathways when comparing to healthy controls but downregulation when comparing to non-lesional controls.

### Identification of disease-specific molecular features

Despite the shared clinical manifestations, Acne vulgaris, PPR, and HS represent distinct diseases with potentially differing etiologies. To gain deeper insights into their molecular distinctions, we conducted a comparative analysis of the DEGs across these three conditions, identifying DEGs that are either exclusive to each disease or commonly shared (**Fig. 2a; Supplementary Fig.2a**). Our approach began by comparing the DEGs lists sourced from the three primary datasets (Acne vulgaris GSE6475, PPR GSE65914, and HS GSE148207), and then validated these genes by cross-referencing them with the additional eight datasets listed in Supplementary Fig. 1a. (e.g., we verified that the DEGs unique to Acne vulgaris did not appear in the DEG lists from all PPR and HS datasets). In the case of PPR, there were 216 uniquely upregulated and 209 uniquely downregulated DEGs. Notably, HS exhibited the most extensive set of exclusive DEGs, encompassing 651 upregulated and 658 downregulated genes. This pattern of distinct DEGs remained consistent across all diseases when comparing lesional samples to non-lesional controls (**Supplementary Fig.2a**).

**Figure 2.**
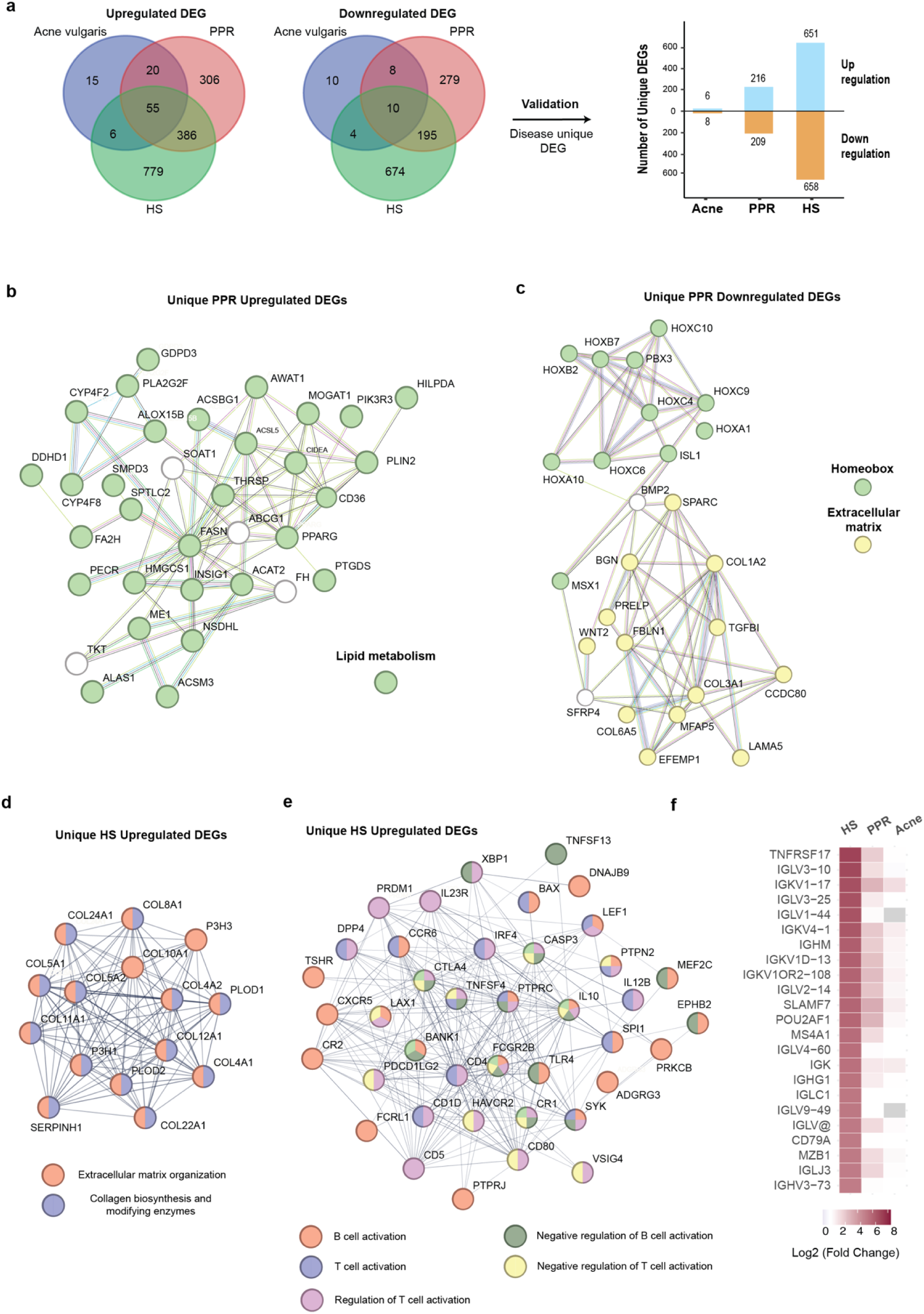
Identification of molecular features that are unique to each disease. **a)** Strategy for identification of unique DEGs. Venn plots were generated by overlaying DEGs from the three diseases, considering upregulated and downregulated DEGs separately. DEGs that were unique to each disease were validated against additional datasets to ensure the uniqueness. The validated unique DEGs were quantified and visualized using a bar graph. **b)** A protein- protein interaction (PPI) network of DEGs from unique PPR upregulated genes that are enriched in the lipid metabolism pathway. **c)** A PPI network of DEGs from unique PPR downregulated genes that are enriched in the Homeobox gene family and the extracellular matrix pathway. **d)** A PPI network of unique HS upregulated genes that are enriched in extracellular matrix organization and collagen biosynthesis and modifying enzymes. **e)** A PPI network of unique HS upregulated genes that are enriched in B cell and T cell activation and regulation pathways. **f)** Upregulation of gene expression in B cell/plasma cell-related genes in HS comparing to PPR and Acne vulgaris.

We then performed STRING protein-protein interaction network analysis on the DEGs that were specific to PPR and HS (**Fig. 2b-f and Supplementary** Fig. 2b-e). When comparing lesional to the healthy controls, PPR displayed uniquely upregulated genes that were enriched in lipid metabolism pathways (**Fig. 2b**). This observation aligns with the GSEA comparison results depicted in Figure 1D, which also highlighted the differential upregulation of lipid metabolism-related pathways in PPR. Intriguingly, we also detected a distinctive downregulation of development related Homeobox family genes in PPR, including HOXA1, HOXA10, HOXB2, HOXB7, HOXC4, HOXC9, HOXC10, and their interacting partners PBX3 and ISL1 (**Fig. 2c**). Within the same downregulated gene set, we also noticed an enrichment in extracellular matrix proteins, such as COL1A2, COL3A1 and COL6A5, which contrasted with an upregulation in extracellular matrix organization pathways in HS (**Fig. 2c and d; Supplementary Fig.2b**). In the context of comparing lesional to non-lesional samples, we identified that PPR displayed unique downregulation of the late cornification envelope (LCE) family genes implicated in keratinization (**Supplementary Fig. 2c**).

HS exhibited a more pronounced activation of adaptive immune responses when compared to PPR and Acne vulgaris, particularly in B cell/plasma cell responses (**Fig. 2e and Supplementary Fig. 2d**). We then scrutinized the original DEG list for HS and discovered that among the top 50 most-upregulated genes (ranked by Log2 fold change), 24 were associated with B cell/plasma cell functions (**Fig. 2f**). These same genes demonstrated relatively lesser upregulation in PPR and displayed minimal or no upregulation in Acne vulgaris. Of interest, we conducted a comparison between non-lesional samples and healthy controls within the HS dataset. Strikingly, we identified an upregulation of B cell-related genes even in non-lesional samples, implying a systemic elevation of B cell response among HS patients (**Supplementary Fig. 2e**).

### Identification of disease-shared molecular features

Acne vulgaris, PPR and HS are all inflammatory skin disorders, and understanding of the commonality of the molecular features of the three diseases may foster the development of broadly applicable therapeutics. We identified the DEGs that were shared by all three diseases as depicted in Figure 2a and validated them by selecting the DEGs that were also present in the DEG lists of the additional eight datasets. Ultimately, we confirmed the validation of 22 upregulated DEGs, including antimicrobial peptides (S100A7, S100A8, S100A9 and S100A12), chemokines (CXCL1, CXCL8 and CCL19), Matrix Metallopeptidase (MMP1 and MMP12), and protease inhibitor (PI3, SERPINA1 and SERPINB4) (**Fig. 3a**). We performed STRING network analysis on these 22 DEGs and the results revealed the genes were enriched in stress response pathways, chemotaxis, IL-17 signaling pathways, metal sequestration antimicrobial proteins, serine-type endopeptidase activity and serine protease inhibitor (**Fig. 3b**). No shared downregulated DEGs passed validation.

**Figure 3.**
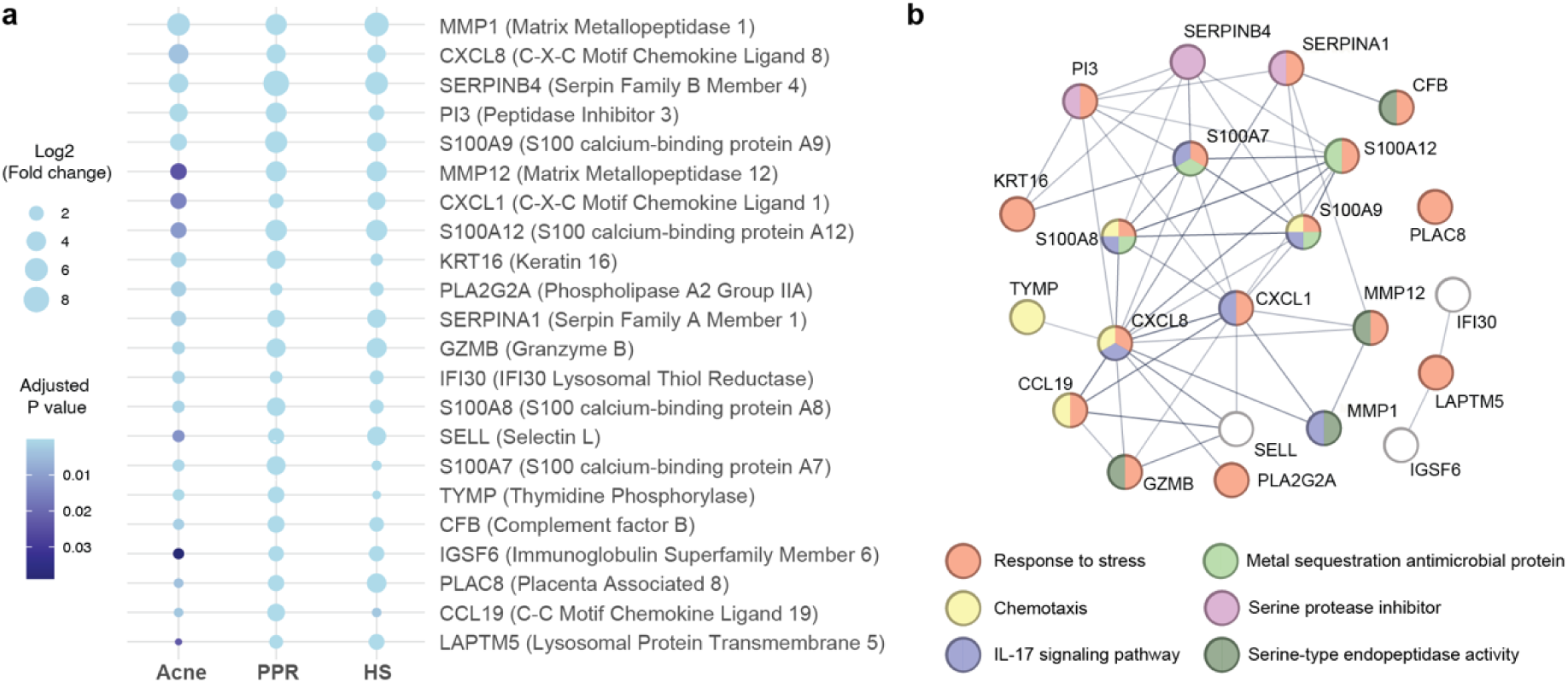
Identification of molecular features that are shared by the three diseases. **a)** Visualization of the 22 validated shared DEGs. The dot size represents the fold change of gene expression, and the dot color scale represents the adjusted P value. **b)** A protein-protein interaction network of the 22 genes. The enriched pathways were highlighted in colors.

### Identification and validation of high dose dietary zinc as a therapeutic agent

To identify potential therapeutic agents, we focused on the shared DEGs and map the gene-drug interactions using the Drugbank database (https://go.drugbank.com/). We observed that multiple genes (SERPINA1, S100A7, S100A8, S100A9 and KRT16) interacted with zinc and its related salts (zinc sulfate, zinc chloride and zinc acetate) (**Fig. 4a**). Zinc is an essential mineral that plays a crucial role in various bodily function, such as immune system regulation. Although zinc supplementation has been explored for the treatment of inflammatory skin disorders, the outcomes have yielded mixed results (Dhaliwal et al. 2020). An examination of the effects of zinc supplementation on inflammatory skin diseases divulged that out of 14 studies focusing on zinc supplementation for acne vulgaris patients, 10 exhibited a beneficial impact on disease severity. Similarly, for HS patients, 3 out of 3 studies demonstrated favorable outcomes from zinc supplementation. However, a study involving rosacea patients failed to demonstrate positive results in comparison to the placebo group. It’s worth noting that these studies have certain limitations, such as varied methodologies with different types of zinc salts and dosages (Dhaliwal et al. 2020). Furthermore, the mechanism underlying zinc’s potential efficacy remains elusive, underscoring the need for further research to comprehensively assess the effectiveness of zinc in treating skin inflammation, especially on the mechanisms underlying the beneficial effects at the molecular level.

**Figure 4.**
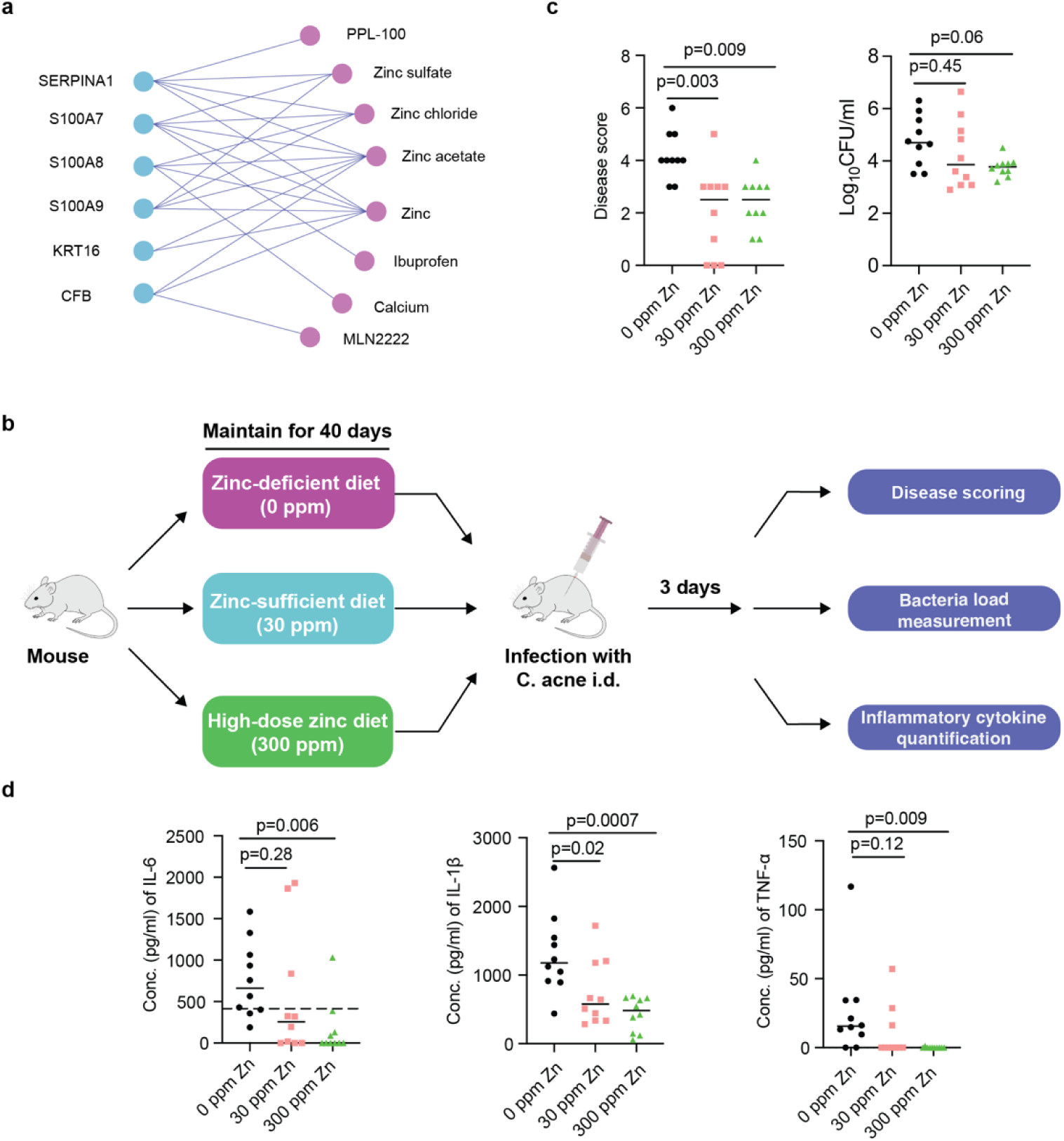
Identification and validation of high dose dietary zinc as a therapeutic agent. **a)** Gene-drug interaction analysis identified zinc and its related salts interacted with multiple validated shared DEGs. **b)** Schematic of the experimental design. **c)** Effect of dietary zinc on disease scores and bacteria load in an acne mouse model that was infected with C. acne HL043PA1 strain. **d)** Quantification of inflammatory cytokines from skin homogenates collected from infected mice under diets with different amount of zinc. All data were presented as medians from two independent experiment (n = 5 for each experiment).

We have previous developed an acne mouse model (Kolar et al. 2019). To directly query the role of zinc in acne, we maintained CD1 mice on a zinc-deficient diet (0 ppm, parts per million) and compared them with the age-matched mice subjected to diets either sufficient in zinc (30 ppm) or featuring a higher zinc concentration (300 ppm) over a 40-day period (**Fig. 4b**). The mice were challenged with C. acnes bacteria intradermally, and disease score and tissue cytokines production were surveyed on Day 3 (**Fig. 4c and d**). Independent of bacterial burden, mice fed on a Zn deficient diet showed significantly higher disease score and inflammation than the other two groups of mice, which showed comparable acne immunopathology. Among the mice fed with zinc, those receiving a high dose (300 ppm) demonstrated more substantial improvements than their counterparts who received a sufficient dose (30 ppm) of zinc.

We tested two C. acnes strains in this study, and notably, we found significant differences in immunopathology with HL043PA1 strain only, while disease score and inflammation were trending upwards in zinc deficient mice when acne was induced with less acnegenic strain HL043PA2 (**Supplementary Fig. 4**). Overall, our findings suggest that dietary zinc could play a significant role in mitigating acne immunopathology by controlling excessive inflammatory phenotype observed in Zn deficient mice.

## Discussion

Acne vulgaris, Rosacea (specifically the Papulopustular Rosacea subtype, PPR), and Hidradenitis Suppurativa (HS) are chronic inflammatory skin disorders with overlapping clinical manifestations. While traditional diagnosis relies on dermatological assessments, accurate prognosis remains challenging due to potential co-existence and progression from one condition to another. The current study leverages omics technologies to elucidate the molecular underpinnings of these conditions. By dissecting three core datasets and complementing our findings with validation from eight additional datasets, we successfully unearthed differentially expressed genes (DEGs) and enriched pathways that are unique to each disorder and collectively shared by all three.

The most significant shared feature of the three diseases is elevated immune and defense responses. Of note, the S100 family proteins, encompassing S100A7, A8, A9, and A12, were consistently upregulated across all three conditions. Intriguingly, enhanced expression of S100 family proteins has also been observed in other inflammatory skin disorders, including psoriasis. Despite their substantial relevance, research and therapeutic endeavors focused on skin inflammatory disorders have largely overlooked this family of proteins. It has been suggested that S100 family proteins can function as antimicrobial peptides but also attract and stimulate immune cell activation (Eckert et al. 2004). Keratin 16 (KRT16) and its partner keratin 6 (KRT6) were also consistently upregulated across the primary datasets for all three diseases. Keratin 16 and 6 are rapidly induced in keratinocytes upon skin injury, and are critical in maintaining keratinocytes’ cell-cell and cell-matrix adhesion (Zhang, Yin, and Zhang 2019). In addition, keratin 16 has been shown to regulate innate immunity in response to epidermal barrier breach (Lessard et al. 2013). Modulating the functions of those immune response regulators presents an intriguing therapeutic option. Through gene-drug interaction analysis, we uncovered a connection between these genes and zinc and its related salts. The beneficial effect of using zinc supplement in treating skin inflammatory disorders is controversial but promising, particularly for acne vulgaris and HS. Our results on murine acne model further validated the benefits of dietary zinc in reducing the disease score and inflammation in a more controlled experimental condition. While the effect of dietary zinc in alleviating disease severity can stem from various factors, its interaction with immune response regulators may wield a crucial influence, particularly considering the pronounced presence of genes within the disease’s molecular features. Intriguingly, our findings also revealed that mice fed a higher dose of zinc exhibited superior disease scores and a reduced quantity of inflammation-related cytokines compared to those mice provided with a merely sufficient amount of zinc. This dose-dependent effect echoes the outcomes observed in a zinc supplement study for HS patients, wherein a reduction in zinc dosage led to disease recurrence (Dhaliwal et al. 2020). Moreover, this insight offers a plausible explanation for the divergent clinical results of zinc supplementation, underscoring the importance of controlling zinc amounts according to a universally applicable protocol.

Comparative transcriptome analysis can also shed light on the distinctiveness of each disease, offering insights into the underlying variations in their etiologies. Intriguingly, our results highlighted HS’s pronounced recruitment and activation of B cell/plasma cells when contrasted with Acne vulgaris and PPR, with nearly half of the top 50 upregulated DEGs being related to B cell/plasma cells. Data from Gudjonsson et al. also revealed that HS skins had increased immunoglobulin production and complement activation (Gudjonsson et al. 2020). More strikingly, we observed an upregulation of B cell/plasma cell activity even in non-lesional skins of HS patients compared to healthy controls, suggesting a systematic immune activation among HS patients that renders their skin more susceptible to lesion formation. It offers a potential rationale for the limited efficacy of Acne vulgaris medication in treating patients with HS. In addition, given the limited therapeutic options for HS, targeting B cell/plasma cell activity might serve as a potent therapeutic avenue. Another intriguing revelation is the unique elevation of collagen gene expression in HS patients comparing to PPR and Acne vulgaris, potentially correlating with heightened fibroblast activation and scar formation. Interestingly, we also observed an association of Homeobox family genes with Rosacea (i.e., PPR downregulated the expression of seven HOX genes), which is not observed in Acne vulgaris and HS. Homeobox family genes are known to be involved in skin development and have been suggested to play a role in wound healing. HOX-B13 is markedly downregulated in response to wounding, and HOXA7/A9/C8 are shown to be significantly lower in keloid fibroblasts comparing to normal fibroblasts (Namazi, Fallahzadeh, and Schwartz 2011; Hahn et al. 2021). Although targeting inflammation is the current mainstream therapeutic strategies in treating skin inflammatory skin disorders, modulating the post-inflammation skin regeneration is also critical to reduce the appearance of permanent scarring.

In summary, we successfully demonstrate the potential of comparative omics analysis in delving into the etiologies of skin disorders characterized by shared clinical manifestations and developing therapeutics based on the understanding of the underlying molecular features. This strategy could evolve to explore the fundamental drivers of skin disorders, guiding the development of ‘root’-targeted therapeutics, diverging from transient immunosuppressive drugs fraught with infection and cancer-related risks. The major challenge we faced during the project is the integration of omics data from different platforms. We propose to establish a skin and skin disorder omics data consortium with community-curated standards as a step toward precision dermatology.

## METHODS

### Microarray and RNA-Seq data collection

Microarray and RNA-Seq studies published until July 2023, focusing on skin samples associated with Rosacea, Acne, and hidradenitis suppurativa, were sourced by querying the Gene Expression Omnibus (GEO) (Barrett et al. 2012) using the following key words: (Acne OR Rosacea OR hidradenitis suppurativa OR HS) AND (RNA-seq OR Expression profiling OR high throughput sequencing). The result was manually filtered that are only from expression profiling by Affymetrix array and Illumina high-throughput sequencing. Raw expression data and accompanying metadata were acquired using the GEOquery package v2.68.0 (Davis and Meltzer 2007). A complete list of studies included in the analysis can be found in Supplementary Figure 1a. Each Sample was manually labeled as “Healthy”, “Non-lesional” or “Lesional”, corresponding to healthy control, non-lesional sample from patients, and lesional sample from patients, respectively.

### Gene expression quantification for microarray data and RNA-seq data

The microarray data were pre-processed through the Bioconductor affy package v1.78.0 in R to generate count matrix with Robust Multi-Array Average (RMA) expression measure (Davis and Meltzer 2007). The data were then annotated by hgu133plus2.db or corresponding annotation table by query the GPL accession number in GEO database (Carlson et al. 2016). The differential expression analysis was then performed by the limma package v3.56.2 in R. The up- and down-regulated differentially expressed genes (DEGs) were selected from each dataset using a threshold of log2 fold change of 1 and an adjusted p–value (Bonferroni correction) of 0.05 (Ritchie et al. 2015). For RNA-seq data, raw counts were initially annotated using Genome assembly GRCh38.p13. Differential expression analysis was subsequently conducted on the raw counts using the DESeq2 package v1.40.2, employing a negative binomial generalized linear model (Love, Huber, and Anders 2014). The same fold change and adjusted p-value thresholds applied to the microarray data analysis were used here as well. Notably, the outcome of differential expression analysis from rosacea dataset 2 was directly acquired from the Mendeley data website. (Shih et al. 2020). The Venn plots were created using tools from https://bioinformatics.psb.ugent.be/webtools/Venn/.

### Gene set enrichment analysis

The analysis was conducted using the R package clusterProfiler v3.12.047. Gene sets and molecular signatures utilized in this analysis were obtained from the Molecular Signatures Database. Enrichment scores were normalized and calculated through 1000 gene set permutations. Subsequently, significant enrichment outcomes were filtered based on a P-value cutoff of 0.05.

### Protein-protein interaction (PPI) network analysis

The STRING database (https://string-db.org) was employed to predict functional interactions among the differentially expressed genes. The network type was designated as a full string network. In the case of HS, a high confidence threshold (0.700) and high FDR stringency (1 percent) were applied. For the remaining cases, a medium confidence threshold (0.400) and medium FDR stringency (5 percent) were used.

### Ethics statement

All mouse studies were approved under the guidelines of the University of California San Diego (UCSD) Institutional Animal Care and Use Committee (S18200). Outbred 6 weeks old female CD1 mice (The Charles River Laboratory) were housed in an animal facility at UCSD with a standard of care as per federal, state, local, and NIH guidelines.

### *C. acnes* bacterial culture

This study involved the utilization of two acne-associated strains, namely HL043PA1 and HL043PA2. As established in our previous investigation [1], these strains are representative of the most acnegenic strains within the ribotype-type 4/5 panel utilized in our acne mouse model (Kolar et al. 2019). The clinical C. acnes strains sourced from frozen stock were cultivated anaerobically on blood agar plates using the BD BBL™ GasPak™ system for a duration of 96 hours at 37 °C. A single colony of *C. acnes* was anaerobically grown in 10 ml of Brain Heart Infusion (BHI) broth (Catalog no. #53286, Sigma-Aldrich, USA) for 3-4 days (OD=0.15-0.3), followed by washing once of the bacterial pellet with BHI media at 2300 × g for 5 minutes. The pellet was resuspended in BHI media to an OD of 0.5 for *in vivo* mouse infection.

### Diets

Six-week-old mice were assigned randomly to three distinct experimental groups. These groups were characterized by their dietary intake: a zinc-deficient diet (egg white-based AIN-93G without added zinc; a diet with a 30 ppm concentration of zinc (egg white-based AIN-93G with added zinc); or a diet with 300 ppm concentration of Zn (egg white-based AIN-93G with 300 ppm added zinc). The diets were procured from Research Diets, Inc., and the diets were irradiated with 10 to 20 kGy gamma-irradiation. All the three groups of mice were provided with sterile food and water ad-libitum, and the mice were maintained on diets for a period of 40 days. The composition details of the diets are provided in Supplementary Table 2.

### Acne mouse model

To model human acne disease, mice that were on different diets were intradermally (i.d.) infected with a *C. acnes* strain (2x10^7^CFU in 50μl volume of BHI media), followed by the topical application of synthetic sebum daily as described previously (Kolar et al. 2019). The i.d. infections were performed under vaporized Isoflurane (Fluriso, Vet One) anesthesia. On the third day post-infection, the disease score was evaluated, and euthanasia was carried out using CO_2._ Skin lesions were aseptically excised and harvested in 400 µl sterile phosphate buffer saline (PBS, pH 7.4). The skin lesions were then homogenized and 25 μl was serially diluted (10-fold) in PBS to determine CFU on BHI agar plates. The BHI agar plates were incubated anaerobically at 37 °C for a period of 3-4 days. In addition, the homogenized skin lesions were centrifuged at maximum speed (13000 rpm) for 25 min, yielding a supernatant that was subsequently stored at -80°C for the purpose of cytokine analysis.

### Disease scoring

The assessment of gross skin pathology involved scoring according to the following criteria: Erythematous change (none = 0, mild = 1, moderate = 2, and marked = 3); and papule characteristics (flat = 0, small = 1, large = 2, and extra-large = 3).

### Determination of cytokines in skin lesions

IL-1β, IL-6, and TNF-α cytokine levels within stored skin homogenates at -80°C were quantified through a solid-phase sandwich ELISA, employing commercially available mouse cytokine ELISA kits (Biolegend, San Diego, CA, USA). For IL-1β and IL-6, skin homogenates (50 μl) were diluted at a 1:1 ratio with blocking buffer (1% BSA plus 1xPBS-Tween20), while undiluted skin homogenate (100 μl) was used for TNF-α. These were assayed alongside cytokine standards of known concentrations (provided with the kits) on each ELISA plate. The plates were developed, and the optical density (OD) was read at 450 nm with wavelength correction set at 570 nm, utilizing a multimode microplate reader (PerkinElmer, Waltham, MA, USA). The cytokine levels in the samples were determined using the standard curve derived from the OD of the cytokine standards.

### Statistical analysis

GraphPad prism version 8 was used to analyze all data (GraphPad Software, San Diego, CA, www.graphpad.com). All mouse model data were presented as medians from two independent experiments. For comparisons involving multiple groups, a one-way ANOVA with Tukey’s post-hoc test was employed (HL043PA1 bacteria load and IL-1β). If normality assumptions were not met, non-parametric Kruskal-Wallis one-way ANOVA was utilized to analyze the data (HL043PA1 disease score, IL-6, and TNF-α).

## Abbreviations

PPR: Papulopastular Rosace
HS: Hidradenitis Suppurativa
DEG: Differentially Expressed Gene
GSEA: Gene Set Enrichment Analysis

## DATA AVAILABILITY

All primary gene expression data are available from Gene Expression Omnibus. All analyzed results are available on Zenodo (8343604).

## DECLARATION OF GENERATIVE AI AND AI-ASSISTED TECHNOLOGIES IN THE WRITING PROCESS

During the preparation of this work the author(s) used ChatGPT 3.5 in order to improve the readability and language of manuscript. After using this tool/service, the author(s) reviewed and edited the content as needed and take(s) full responsibility for the content of the publication.

## AUTHOR CONTRIBUTION

Conceptualization: LL, JM, YW, GMC, GL Data Curation: LL, ZT, YW, IH

Formal Analysis: LL, ZT, YW, IH Funding Acquisition: LL, YW, GMC, GL

Investigation: LL, ZT, YW, IH, NB, ZZ Supervision: GMC, YW, GL, JM Visualization: LL, ZT, YW, IH, YZ Writing – YW

Writing - Review and Editing: ALL

## ACKNOWLEDGEMENT

Portions of this research were conducted on the O2 High Performance Compute Cluster, supported by the Research Computing Group, at Harvard Medical School. This work was supported by Leo Foundation (LF-OC-20-000420), Strategic Priority Research Program of The Chinese Academy of Sciences (XDB0480000) and National Institute of Health (R01AI141401).

