## Supplementary materials for "Comparative Transcriptome Analysis of Acne vulgaris, Rosacea, and Hidradenitis Suppurativa Supports High Dose Dietary Zinc as a Therapeutic Agent" (from another version of this preprint: version 1, 2023-11-29)

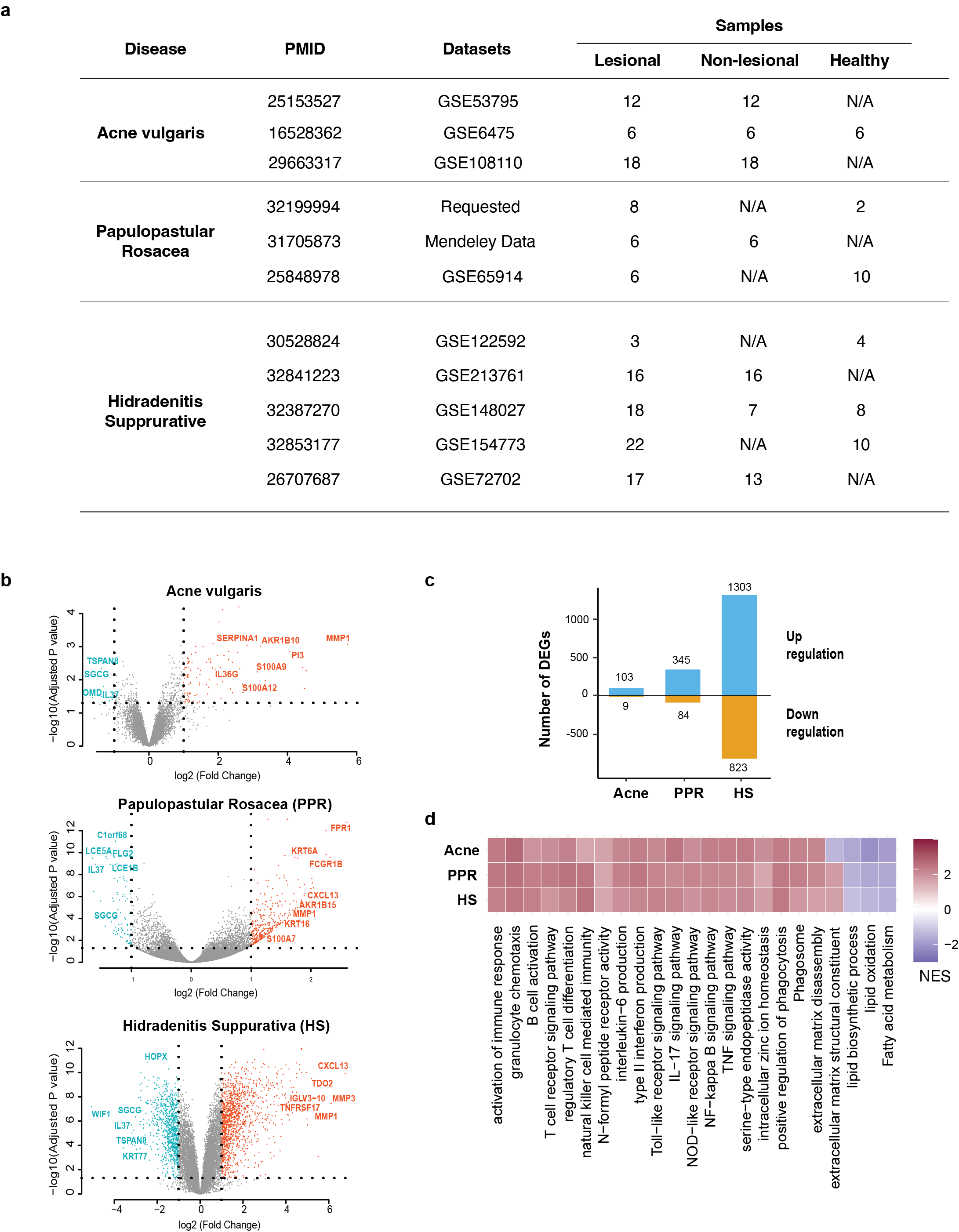

**Supplementary figure 1. Transcriptome analysis of Acne vulgaris, Papulopastular rosacea and Hidradenitis suppurativa (lesional versus non-lesional controls). a)** Description of datasets used in the project. **b)** Volcano plots illustrating the distribution of differentially expressed genes for the three diseases. **c)** Summary of the number of DEGs for the three diseases. **d)** Selected enriched pathways from gene set enrichment analysis for each disease. NES: normalized enrichment score.

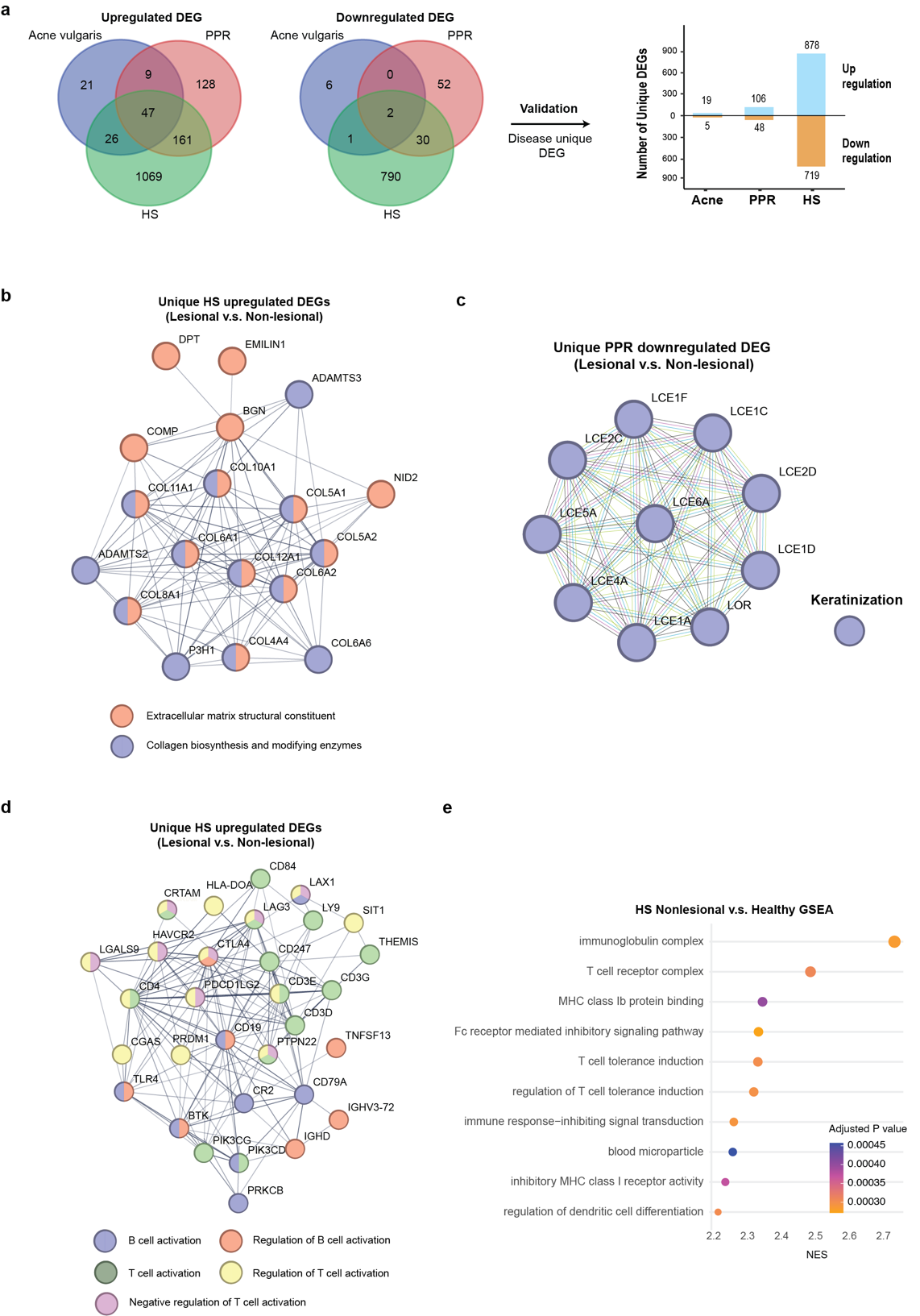

**Supplementary figure 2.** **Identification of molecular features that are unique to each disease (lesional versus non-lesional controls). a)** Strategy for identification of unique DEGs for each disease. Venn plots were generated by overlaying DEGs from the three diseases, considering upregulated and downregulated DEGs separately. DEGs that were unique to each disease were validated against additional datasets to ensure the uniqueness. The validated unique DEGs were quantified and visualized using a bar graph. **b)** A PPI network of unique HS upregulated genes that are enriched in extracellular matrix organization and collagen biosynthesis and modifying enzymes. **c)** A protein-protein interaction (PPI) network of DEGs from unique PPR upregulated genes that are enriched in the keratinization. **d)** A PPI network of unique HS upregulated genes that are enriched in B cell and T cell activation and regulation pathways. **e)** Activation of immune response in HS patient non-lesional samples comparing to healthy controls.

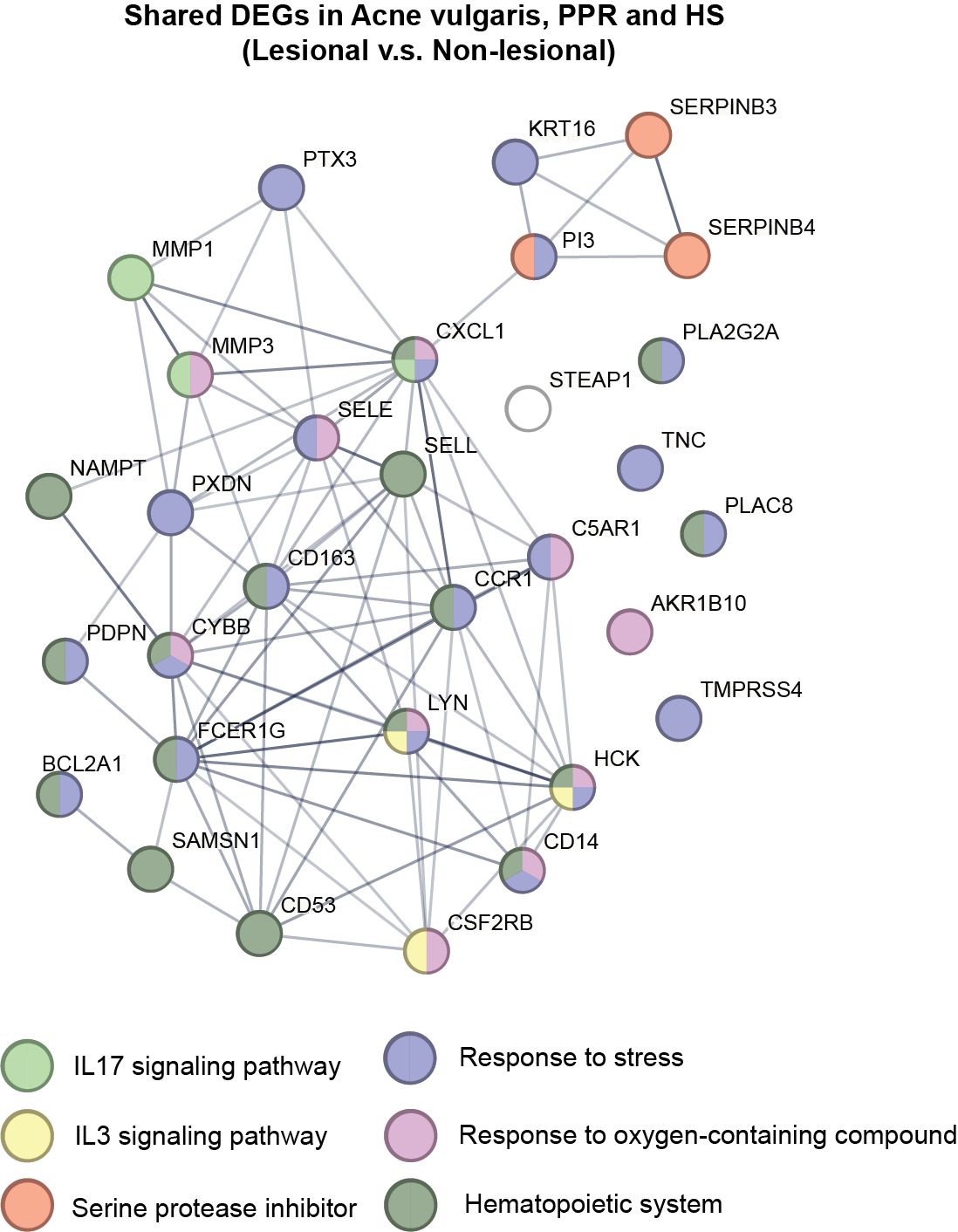

**Supplementary figure 3.** **Identification of molecular features that are shared by the three diseases (lesional versus non-lesional controls).** Overlapped DEGs from the three core datasets were validated by cross-referencing the supplementary datasets. A protein-protein interaction network of the validated 31 upregulated genes was displayed. The enriched pathways were highlighted in colors.

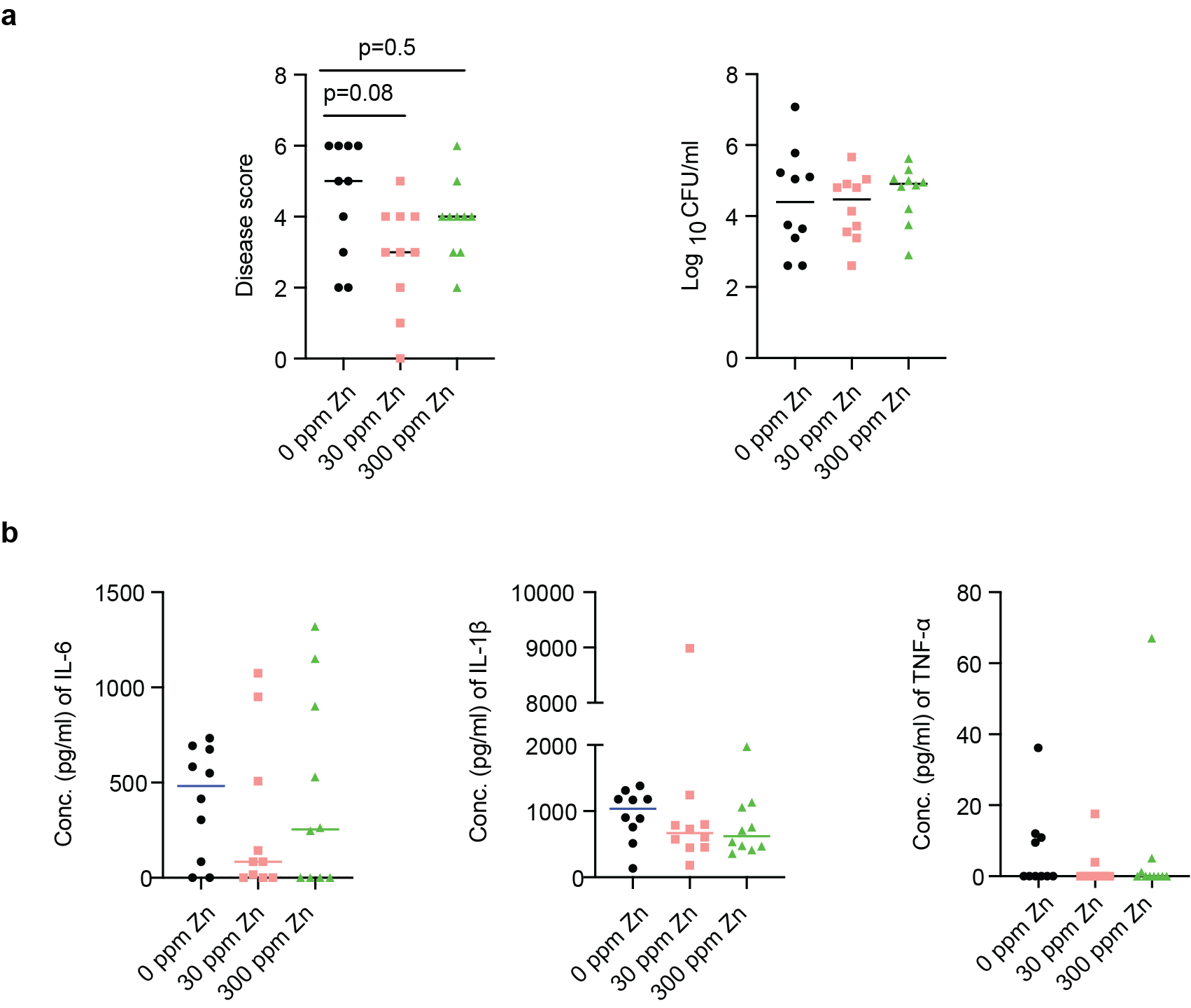

**Supplementary figure 4. Test of high dose dietary zinc on an acne mouse model infected with less pathogenic C. acne HL043PA2 strain. a)** Effect of dietary zinc on the disease score and bacteria load in an acne mouse model that was infected with C. acne HL043PA2 strain. **b)** Quantification of inflammatory cytokines from skin homogenates collected from infected mice under diets with different amount of zinc. All data were presented as medians from two independent experiment (n = 5 for each experiment).

**Supplementary Table 1. Gene-drug interaction analysis for disease-shared DEGs.**

| **Gene.Name** | **Drug.Names** |
| --- | --- |
| CFB | Zinc |
| CFB | 4-guanidinobenzoic acid |
| CFB | Diisopropylphosphono Group |
| CFB | MLN2222 |
| CFB | Zinc acetate |
| CXCL8 | ABT-510 |
| CXCL8 | MDX-018 |
| CXCL8 | Rivanicline |
| CXCL8 | Tapinarof |
| KRT16 | Zinc |
| KRT16 | Zinc acetate |
| MMP1 | Marimastat |
| MMP1 | CGS-27023 |
| MMP1 | N-[3-(N'-Hydroxycarboxamido)-2-(2-Methylpropyl)-Propanoyl]-O-  Tyrosine-N-Methylamide |
| MMP1 | Methylamino-Phenylalanyl-Leucyl-Hydroxamic acid |
| MMP1 | [[1-[N-Hydroxy-Acetamidyl]-3-Methyl-Butyl]-Carbonyl-Leucinyl]-  Alanine Ethyl Ester |
| MMP1 | N-Hydroxy-2-[4-(4-Phenoxy-Benzenesulfonyl)-Tetrahydro-Pyran-4-YL]-  Acetamide |
| MMP12 | Acetohydroxamic acid |
| MMP12 | Marimastat |
| MMP12 | CP-271485 |
| MMP12 | PF-00356231 |
| MMP12 | Batimastat |
| MMP12 | 2-[2-(1,3-Dioxo-1,3-Dihydro-2h-Isoindol-2-Yl)Ethyl]-4-  (4'-Ethoxy-1,1'-Biphenyl-4-Yl)-4-Oxobutanoic Acid |
| MMP12 | AE-941 |
| MMP12 | (1S,5S,7R)-N~7~-(Biphenyl-4-YLMethyl)-N~3~-Hydroxy-6,8-DIOXA-3-  Azabicyclo[3.2.1]Octane-3,7-DICARBOXAMIDE |
| MMP12 | N-(biphenyl-4-ylsulfonyl)-D-leucine |
| MMP12 | CGS-27023 |
| MMP12 | N-(dibenzo[b,d]thiophen-3-ylsulfonyl)-L-valine |
| MMP12 | N-oxo-2-[(4-phenylphenyl)sulfonylamino]ethanamide |
| MMP12 | 2-[(4-fluorophenyl)sulfonylamino]-N-oxo-ethanamide |
| MMP12 | N-oxo-2-(phenylsulfonylamino)ethanamide |
| MMP12 | N-Isobutyl-N-[4-Methoxyphenylsulfonyl]Glycyl Hydroxamic acid |
| MMP12 | N-[(4-methoxyphenyl)sulfonyl]-D-alanine |
| PLA2G2A | Indomethacin |
| PLA2G2A | Ginkgo biloba |
| PLA2G2A | 1,4-Butanediol |
| PLA2G2A | 1-{2-[2-(2-Methoxyethoxy)Ethoxy]Ethoxy}-4-(1,1,3,3-  Tetramethylbutyl)Benzene |
| PLA2G2A | Isopropyl alcohol |
| PLA2G2A | N-Tridecanoic Acid |
| PLA2G2A | [3-(1-Benzyl-3-Carbamoylmethyl-2-Methyl-1h-Indol-5-Yloxy)-  Propyl-]-Phosphonic Acid |
| PLA2G2A | 4-(1-Benzyl-3-Carbamoylmethyl-2-Methyl-1h-Indol-5-Yloxy)-  Butyric Acid |
| PLA2G2A | Lauric acid |
| PLA2G2A | (1-Benzyl-5-methoxy-2-methyl-1H-indol-3-yl)acetic acid |
| PLA2G2A | 6-phenyl-4(R)-(7-phenyl-heptanoylamino)-hexanoic acid |
| PLA2G2A | Elaidamide |
| PLA2G2A | (2R)-2-Acetamido-3-(octadecyloxy)propyl 2-  (methylsulfanyl)ethyl hydrogen phosphate |
| PLA2G2A | KH064 |
| PLA2G2A | Suramin |
| PLA2G2A | Varespladib methyl |
| S100A12 | Olopatadine |
| S100A12 | Amlexanox |
| S100A7 | Ibuprofen |
| S100A7 | Zinc |
| S100A7 | 8-anilinonaphthalene-1-sulfonic acid |
| S100A7 | Dexibuprofen |
| S100A7 | Zinc acetate |
| S100A7 | Zinc chloride |
| S100A7 | Zinc sulfate, unspecified form |
| S100A8 | Calcium |
| S100A8 | Zinc |
| S100A8 | Copper |
| S100A8 | Zinc acetate |
| S100A8 | Zinc chloride |
| S100A8 | Zinc sulfate, unspecified form |
| S100A9 | Calcium |
| S100A9 | Zinc |
| S100A9 | Zinc acetate |
| S100A9 | Zinc chloride |
| S100A9 | Zinc sulfate, unspecified form |
| SERPINA1 | Zinc |
| SERPINA1 | 2-[3,4-Dihydroxy-2-Hydroxymethyl-5-(2-Hydroxy-Nonyl)-Tetrahydro-  Furan-2-Yloxy]-6-Hydroxymethyl-Tetra Hydro-Pyran-3,4,5-Triol |
| SERPINA1 | Mercaptoethanol |
| SERPINA1 | Recombinant alpha 1-antitrypsin |
| SERPINA1 | PPL-100 |
| SERPINA1 | Copper |
| SERPINA1 | Zinc acetate |
| SERPINA1 | Zinc chloride |
| SERPINA1 | Zinc sulfate, unspecified form |
| TYMP | Tipiracil |

**Supplementary Table 2. Composition details of the diets used in this study**

|  | **Zinc-deficient diet** | | **Zinc-sufficient diet** | | **High dose zinc diet** | |
| --- | --- | --- | --- | --- | --- | --- |
|  | gm% | *kcal%* | gm% | *kcal%* | gm% | *kcal%* |
| Protein | 20 | *20* | 20 | *20* | 20 | *20* |
| Carbohydrate | 64 | *64* | 64 | *64* | 64 | *64* |
| Fat | 7 | *16* | 7 | *16* | 7 | *16* |
| Total |  | *100* |  | *100* |  | *100* |
| kcal/gm | 4.0 |  | 4.0 |  | 4.0 |  |
| **Ingredient** | **gm** | ***kcal*** | **gm** | ***kcal*** | **gm** | ***kcal*** |
| Casein | 0 | *0* | 0 | *0* | 0 | *0* |
| L-Cystine | 0 | *0* | 0 | *0* | 0 | *0* |
| Egg White, Spray Dried | 203 | *812* | 203 | *812* | 203 | *812* |
| Corn Starch | 397.486 | *1590* | 397.486 | *1590* | 397.486 | *1590* |
| Maltodextrin 10 | 132 | *528* | 132 | *528* | 132 | *528* |
| Sucrose | 100 | *400* | 100 | *400* | 100 | *400* |
| Cellulose | 50 | *0* | 50 | *0* | 50 | *0* |
| Soybean Oil | 70 | *630* | 70 | *630* | 70 | *630* |
| t-Butylhydroquinone | 0.014 | *0* | 0.014 | *0* | 0.014 | *0* |
| Mineral Mix S10022G | 0 | *0* | 0 | *0* | 0 | *0* |
| Mineral Mix S19409 (no added Ca, P, K or Zn) | 7 | *0* | 7 | *0* | 7 | *0* |
| Potassium Phosphate, Monobasic | 6.86 | *0* | 6.86 | *0* | 6.86 | *0* |
| Calcium Carbonate | 8 | *0* | 8 | *0* | 8 | *0* |
| Potassium Citrate, 1 H2O | 2.4773 | *0* | 2.4773 | *0* | 2.4773 | *0* |
| Calcium Phosphate, Dibasic | 6 | *0* | 6 | *0* | 6 | *0* |
| Zinc Carbonate (52.1% Zinc) | 0 | *0* | 0.0578 | *0* | 0.5737 | *0* |
| Vitamin Mix V10037 | 10 | *40* | 10 | *40* | 10 | *40* |
| Biotin, 1% | 0.4 | *0* | 0.4 | *0* | 0.4 | *0* |
| Choline Bitartrate | 2.5 | *0* | 2.5 | *0* | 2.5 | *0* |
| FD&C Yellow Dye #5 | 0 | *0* | 0 | *0* | 0 | *0* |
| FD&C Red Dye #40 | 0 | *0* | 0 | *0* | 0 | *0* |
| Dye FD&C Blue # 1 | 0 | *0* | 0.05 | *0* | 0 | *0* |
| **Total** | **995.737** | ***4000*** | **995.845** | ***4000*** | **996.311** | ***4000*** |
| Ca (%) | 4.97 |  | 4.97 |  | 4.97 |  |
| P (%) | 3.16 |  | 3.16 |  | 3.16 |  |
| K (%) | 5.15 |  | 5.15 |  | 5.15 |  |
| Zn (mg/kg diet or ppm) | 0 |  | 30 |  | 300 |  |
